# Epidemiological Mapping of Onchocerciasis Hypoendemic Area of North Achefer District, Amhara Regional State, Ethiopia

**DOI:** 10.1101/2022.06.15.22276425

**Authors:** Wuletaw Tadesse Mekonnin, Tadesse Kebede, Sindew Mekasha

## Abstract

**Background:** Onchocerciasis (river blindness) is caused by a filarial nematode worm called *Onchocerca volvulus* encapsulated in nodules under skin. The adult worm logged itself in nodules of cutaneous skin producing thousands of microfilariae per-day those migrating under the dermis of the skin causing cutaneous and eye disease. Ethiopia is one of countries with a high disease burden of onchocerciasis in Africa. Epidemiological mapping of onchocerciasis in hypoendemic area is a first step in elimination programme and to identify intervention eligible areas. Many districts are uncertain about onchocerciasis transmission; in identifying intervention eligible areas especially North Achefer is located adjacent to onchocerciasis endemic district (Alefa) which receiving semi-annual MDA, and its transmission status of the district is not well studied.

**Objective:** The purpose of this study was to assess the epidemiological status of onchocerciasis in the hypoendemic area of the North Achefer district of the Amhara Regional State.

**Methodology:** community-based cross-sectional study design conducted from July to August 2021. Parasitological, immunological and serological (ELISA and RDT) data were collected from the field and for central laboratory. Statistical analysis was conducted using Epi-info software version 7, transported to SPSS software version 26. Descriptive analysis was conducted and presented with frequencies and percentages. The association between dependent and independent variables was analyzed using bivariate logistic regression and variables those with a P-value <0.05% was considered statistically significant.

**Result:** a total of 264 participants enrolled in the study out of which 56.4% were male, with mean age of 28 years. The microscopic examination of skin snip was no microfilariae positive cases, whereas, 3% and 9.1% positive were recorded for Ov16 RDT and Ov16 ELISA test, respectively. Onchocerciasis morbidity indicators were 6 (2.3%), 12 (4.5%), and 9 (3.4%) palpable nodule, skin discoloration, and skin depigmentation respectively. Age, gender, village type, and distance from the river were independent variables that had a significant association with positivity for Ov16 ELISA test.

**Conclusion:** a high prevalence of onchocerciasis exposure which is above the WHO recommended threshold (5%) by Ov16 ELISA assay was observed. Distance from the river, village type, age, and gender had significantly associated with Ov16 ELISA test. Therefore, onchocerciasis elimination intervention in North Achefer is recommended to be implemented. Increasing the sample size and including molecular (i.e. PCR) and entomological technique best recommended to maximize the positivity of the disease and to insure the exact status of the disease in the district respectively.

## Background

Onchocerciasis is one of the Neglected Tropical Disease (NTD) caused by vector-borne filarial nematode *Onchocerca volvulus* (1), transmitted from person to person by the biting of blood feeding female blackfly *Simulium species* (2). Infection by *Onchocerca volvulus* is characterized by itching of the skin and severe eye lesion that leads to long-lasting blindness. The blackfly breed in fast-flowing rivers and streams, hence, the disease onchocerciasis is called “river blindness” (3). The fertilized adult female of *Onchocerca volvulus* worm (which dwells in subcutaneous nodule) produces many thousands of microfilaria per a day that migrate under the skin dermis. The microfilaria freely moves in the intracellular space of the skin causing cutaneous skin lesions; onchodermatitis. The migrating microfilaria invades the conjunctiva and cornea causing inflammation of the eye which results in blindness (4, 5).

Onchocerciasis is the second most disease causing blindness next to trachoma worldwide (6, 7). Onchocerciasis is a major public health problem in six Latin Americans (Colombia, Ecuador, Venezuela, Brazil, Guatemala, and Mexico), Yemen, and 31 Tropical African countries (Nigeria, Senegal, Ethiopia, Sudan, DRC, Kenya… (8). The Latin American countries Colombia, Ecuador and Mexico have eliminated transmission and verified by WHO in 2013, 2014, and 2015 respectively (3). Before onchocerciasis elimination programs launched; over 40 million people were infected and 160 million were at risk of the infection worldwide and more than 99% live in Africa (9).

The first onchocerciasis report in Ethiopia was in 1939 by Italian investigator in Bonga, in Southern Ethiopia (10, 11). In Ethiopia, two species of blackfly vectors are recognized as causing the disease; those are *Simulium damnosum* and *Simulium neavei* (12). The disease is associated with coffee farming in the Southwest and seed oil cultivation in Northwest of the country. About 84% of case is in the Southwestern Ethiopia (13) and by 1997-2001, 2011 and 2012 a Rapid Epidemiological Mapping of Onchocerciasis (REMO) was conducted and confirmed in Northwestern zones of Amhara, Western zones of Oromia, and SNNR, whole Benishangul Gumuz and Gambella, but the disease is 0% in Eastern Ethiopia (14).

In Ethiopia, over 23 million people are at risk of infection by onchocerciasis in surveyed endemic areas (15), and many districts are uncertain for onchocerciasis transmission (16) out which North Achefer district is found in Amhara region, West Gojjam zone. Areas those were not ever assessed and untreated; including hypoendemic areas suitable for onchocerciasis transmission need to be assessed to decide whether a transmission is active or intervention is required. Residents of North Achefer district are suffering from skin pathology, economic, and social impacts due to onchocerciasis infection. Since, the district is bordered on the west by *O. volvulus* endemic Alefa district, there might be a cross-border transmission, and people migrate to the district or the blackfly can move up to 150Km. A survey done by FMoH in 2011 indicates a 17% nodule rate in North Achefer district for onchocerciasis infection, which was not targeted for MDA treatment. In June 2001 Ethiopian government launched a nationwide ivermectin treatment in onchocerciasis endemic areas aiming to achieve 2025 goal of complete elimination of the disease. FMoH in collaboration with other partner organizations, where the annual progress is monitored by the Ethiopian Onchocerciasis Elimination Expert Advisory Committee (EOEEAC); the committee since then gives a technical guidance for the program (17, 18).

Despite Ethiopia launched a nationwide onchocerciasis elimination program with the goal of elimination by 2025, the transmission status of the disease in all parts of hypoendemic areas wasn’t fully ascertained. During the control program of onchocerciasis, treatments were not allowed with <40% onchocerciasis microfiladermia or <20% nodule rate. Areas that were not ever assessed and untreated; including hypoendemic areas suitable for onchocerciasis transmission need to be assessed to decide whether transmission is active or intervention is required (19). North Achefer district is one of the hypoendemic districts for onchocerciasis transmission, but the current transmission status is not well known and elimination program is not undergoing. Therefore, the goal of this study is to map onchocerciasis in North Achefer hypoendemic district based on serological and skin snip data. Thus, as North Achefer district is found adjacent to the area where MDA is launched (Alefa district), so mapping the district for presence of transmission of the disease is an indispensable part to show the prevalence of the disease in the district. This study will help to generate convincing evidence for decision makers.

## Methods and Materials

### Study Area and Population

A community-based cross-sectional study design was conducted using quantitative data collection method among residents of North Achefer district from July 2021 to August 2021 who live along fast-flowing rivers. North Achefer district is found in Amhara regional state West Gojjam zone. The district capital is Liben, located at a distance of 101Km and 591Km far from Bahirdar and Addis Ababa respectively. The district is surrounded by Bahirdar Zuria on East, North Gonder on the West, Lake Tana on the North, South Achefer on the South and Mecha district on the Southeast (Fig. 1). There are many fast-flowing rivers in the district Beles, Beles-outlet, Merfi, Amoragedel, Jihana, Tikur-wuha, Akusti and others are among known rivers in the district those are favorable for the breeding of the blackflies. The total population is estimated to 259,000 (source: North Achefer district health bureau).

**Fig. 1.**
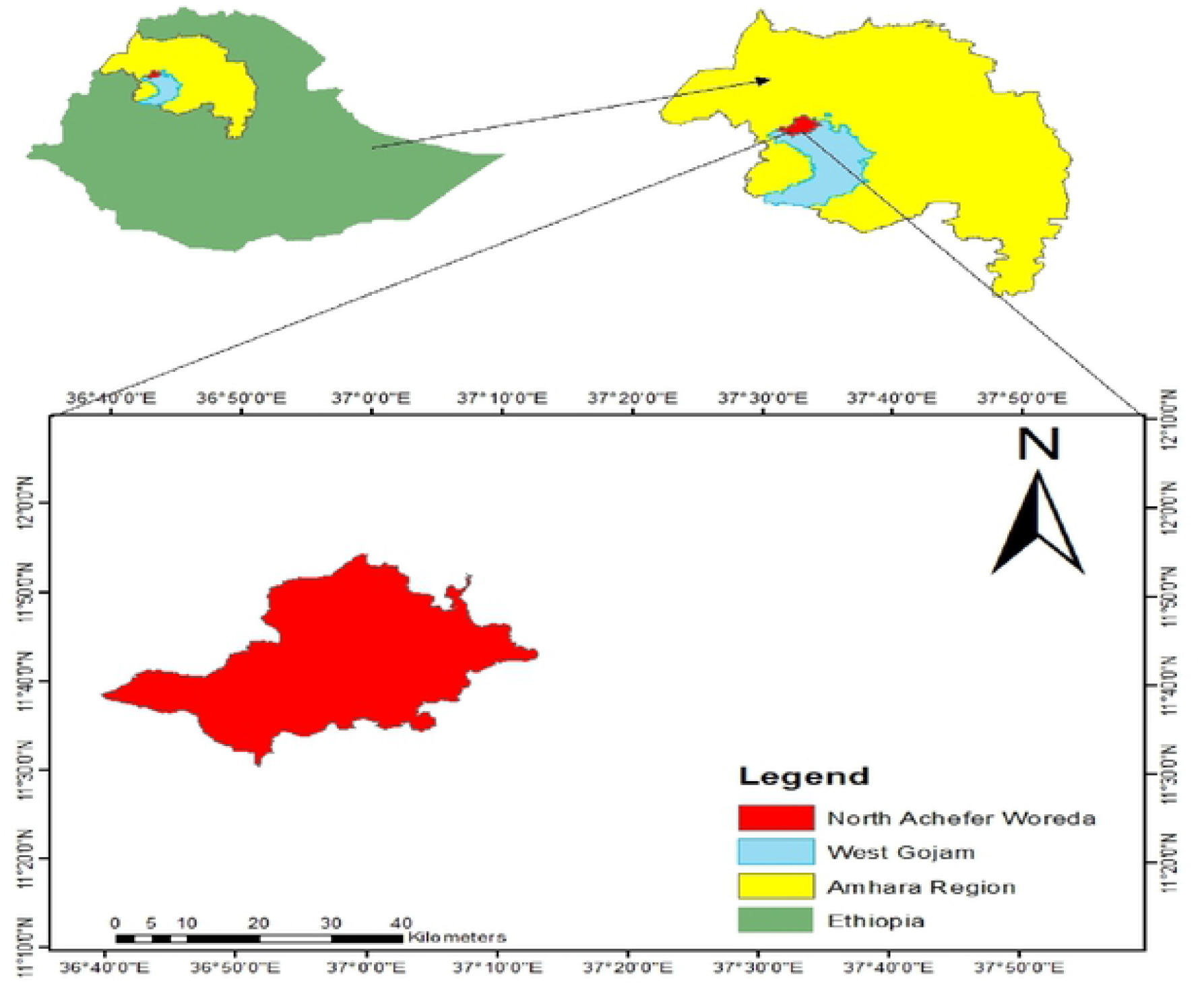
Geographic location of study district, the study map was produced using ArcGIS software version 10.8.

### Sampling Technique and Data Collection

The data was collected from first-line and random village community. Data from the first-line village was collected using local knowledge, proximity to the river, known breeding site, biting complain of the community. Participants of the community were selected using simple random sampling method. A single population proportion formula was used to calculate the total sample size by considering 1.96 for the standard normal variable with 5% level of significance (α-value), 95% confidence interval, 5% margin of error, 10% non-response rate. A prevalence of 19.4% serological test was taken as a reference from a study done in neighboring region Benishangul-Gumuz, Asosa district (20). Therefore, the initial sample size was 240 and with 10% non-response rate, the total sample size was 264. Four villages (Laycheba, Jihana, Dusman and Beles) were selected using purposive sampling method based on proximity to the river, known breeding site, community complain and local knowledge and from two random village (Kunfafila and Beles-outlet). Simple random sampling method was used to select participants of the study; those were ≥5 years living in the village.

The data was collected by volunteer trained health personnel who took the training on the objective of the study with practical scenarios and by principal investigators. After we obtained approval to collect the data from the community leader, the kebele leaders and health extension workers had mobilized the community to gather at the center. All necessary information about the purpose, procedure, and goal of the study was explained for the community, and then data had collected conducted by face-to-face interviews for socio-demographic data, physical examination for morbidity indices, finger prick for the serological test, and skin snip to *O*.*volvulus* microfilaria microscopic examination. All the interviewed questionnaires were checked visually checked by principal investigator and the data coded, cleaned and entered to Epi-info software version 7. Double entry was made to cross-check the data for completeness before analysis. The entered data was exported to SPSS software version 26. Descriptive analysis was done in frequencies, percentages, standard variations and tables. A bivariate analysis was performed to see the frequency distribution and to test whether there is an association between dependent and independent variables respectively. A variables with a P-value of <0.05 declared as having significant statistical association with the outcome variable.

Ethical clearance was received from Addis Ababa University Tikur Anbessa Health Science College Ethical Review Committee and approval was received from Amhara regional State West Gojjam zone health bureau, and North Achefer district administration health office to collect the data. The goal and aim of the study of was explained to the participants and written informed consent was obtained for children’s under 10, from their parents/custodians. Coded ID number was assigned to every participant of the study, which was used only for sample tracking. Original documents and master lists were stored under lock and key in the PI office, which disclosed only for those key personnel who participated in the study.

## Result

### Socio-demographic characteristic of the study participant

This study was done in North Achefer district with a total participants of 264, more than half were male (56.4%) and the mean age of the respondents was 28 years. More than half of the participants were married (57.2%). Out of all respondents, 131 (49.6%) had no formal education, 119 (45.1%) had primary education and 14 (5.3%) were secondary and above. Occupationally, 142 (53.8%) of them were farmers, one (0.4%) was merchant, 16 (6.1%) were kids and 105 (39.7%) were students (Table 1).

**Table 1.** Socio-demographic characteristics of study participants of North Achefer woreda Amhara Region; Ethiopia, June 2022.

| Variable | No of Participant | Percentage |
| --- | --- | --- |
| **Gender** |  |  |
| Male | 149 | 56.4 |
| Female | 115 | 43.6 |
| **Age** |  |  |
| 5 to 10 | 67 | 25.4 |
| 11 to 20 | 57 | 21.6 |
| 21 to 40 | 74 | 27.3 |
| ≥41 | 66 | 25.8 |
| **Marital Status** |  |  |
| Married | 151 | 57.2 |
| Unmarried | 113 | 42.8 |
| **Education** |  |  |
| No formal education | 131 | 49.6 |
| Primary education | 119 | 45.1 |
| Secondary and above | 14 | 5.3 |
| **Occupation** |  |  |
| Farmer | 142 | 53.8 |
| Merchant | 1 | 0.4 |
| Student | 121 | 45.8 |

### Prevalence of onchocerciasis in first-line and random village of North Achefer using ELISA and RDT test

ELISA and RDT serological tests were done; from a total of 264 laboratory test, 8 (4.88) samples ((4(3.8%) from the random village and 4(2.5%)) from the first-line village)) were positive for RDT test and 24 (9.1%) were positive by ELISA test. The ELISA tests from the random village and first-line were 7 (6.7%) and 17 (10.6%) respectively (Table 2).

**Table 2.** Prevalence of onchocerciasis in the first-line and random village of North Achefer Amhara Region; Ethiopia, June 2022.

| Village type |  | OV16 RDT Test Result |  | OV16 ELISA Test Result |  |
| --- | --- | --- | --- | --- | --- |
|  |  | Freq | % | Frequency | % |
| Random | Neg | 100 | 96.2 | 97 | 93.3 |
|  | Pos | 4 | 3.8 | 7 | 6.7 |
|  | Total | 104 | 100.0 | 104 | 100.0 |
| First-line | Neg | 156 | 97.5 | 143 | 89.54 |
|  | Pos | 4 | 2.5 | 17 | 10.6 |
|  | Total | 160 | 100.0 | 160 | 100.0 |

### Prevalence of onchocerciasis morbidity indices in the community of North Achefer woreda

All participants of the study were negative for microfilaria skin snip microscopic examination. As indicated in (Table 3) clinical examination of respondents for onchocerciasis infection morbidity indicators showed that palpable nodule 6 (2.3%), skin discoloration 12 (4.5%), and skin depigmentation 9 (3.4%).

**Table 3.** Prevalence of morbidity indicators of onchocerciasis in a community of North Achefer woreda Amhara region; Ethiopia, June 2022.

| Character | Number | Percentage |
| --- | --- | --- |
| Palpable nodule |  |  |
| Yes | 6 | 2.3 |
| No | 258 | 97.7 |
| Skin discoloration |  |  |
| Yes | 12 | 4.5 |
| No | 252 | 95.5 |
| Skin depigmentation |  |  |
| Yes | 9 | 3.4 |
| No | 255 | 96.6 |

### Knowledge, attitude, and practice of the community for onchocerciasis in North Achefer district

In this study, more than half 163 (61.7%) of the participants have prior information (have heard) about the disease onchocerciasis. Out of 264 respondents, about 123 (46.6%) believe that onchocerciasis is not transmittable disease and only 7 (2.7%) of the study participants knew the disease is caused by filarial nematode. The respondents 89 (33.7%), 111 (42%), 161 (61%) and 80 (30.3%) have believed that onchocerciasis is transmitted by mosquito biting, house-fly biting, contact with an infected person, and aerosol respectively. Nearly half of the respondents 125 (47.3%) knew that the disease is transmitted by the biting of blackfly. Yet, about 80.7% of the study participants reflect their idea as the disease can be eliminated and 88.3% of them answered as they isolate themselves from an infected person by onchocerciasis (Table 4).

**Table 4.** Knowledge, attitude, and practice of the community in North Achefer district Amhara regional state; June 2022.

| Variable's |  | Response (Yes (%)) |
| --- | --- | --- |
| Have you ever heard about onchocerciasis |  | 163 (61.7) |
| Do you think onchocerciasis is a transmittable disease |  | 141 (53.4) |
| Do you know that onchocerciasis cause a filarial worm |  | 7 (2.7) |
| Onchocerciasis is transmitted by | Mosquito biting | 83 (33.7) |
|  | House fly biting | 111 (42) |
|  | Contact with infected person | 161 (61) |
|  | Aerosol | 80 (30.3) |
|  | Black fly biting | 125 (47.3) |
| Do you know how to prevent onchocerciasis |  | 102 (38.6) |

### Factors Associated with Positivity for Onchocerciasis Test

A bivariate logistic regression analysis was done to detect the association between positivity for onchocerciasis and predictive variables for all independent variables. A total of four variables namely distance from the river, sex, village type and age were computed for bivariate logistic regression analysis with a P-value of <0.05. All four variables were indicated a significant association with positivity for Ov16 ELISA test with a P-value <0.05 (Table 5).

**Table 5.** Binary logistic regression analysis result showing factors associated with positivity for OV16 ELISA for onchocerciasis in North Achefer district Amhara region; June 2022.

| Variables |  | OV16 ELISA test |  | OR (95% CI) | P-value |
| --- | --- | --- | --- | --- | --- |
|  |  | Pos | Neg |  |  |
| Sex | Female | 4 | 111 | 0.036 |  |
|  | Male | 20 | 129 | 4.302 (1.428-11.965) | 0.01* |
| Age | 5-10 | 1 | 66 | 0.251 ( 1.26-2.159) | 0.201 |
|  | 11-20 | 6 | 51 | 0.564 (1.099-3.198) | 0.517 |
|  | 21-40 | 11 | 61 | 4.623 (1.492- 14.556) | 0.019* |
| | $\geq 40$ | 6 | 62 | 0.65 | |
| Occupation | Farmer | 13 | 129 | 0.043 (0.06-0.324) | 0.062 |
|  | Student | 11 | 110 | 0.398 (0.09-2.17) | 1 |
|  | Merchant | 0 | 1 | 0.078 |  |
| Village | Random | 7 | 97 | 0.119 | 0.061 |
|  | First-line | 17 | 143 | 3.571 (1.184-10.77) | 0.024* |
| Educ-level | No formal educ. | 14 | 117 | 0.556 (0.189-12.807) | 0.681 |
|  | Prim educ. | 9 | 110 | 0.064 (0.125-9.081) | 0.995 |
|  | Sec. and above | 1 | 13 | 0.77 | 0.134 |
| Dist-(Km) | 0-2 | 11 | 22 | 7.333 (2.725-19732) | 0.008* |
|  | 3-5 | 4 | 86 | 0.682 (0.204-2.285) | 0.535 |
| | $>5$ | 9 | 132 | 0.068 | 0.422 |

Key: ‘\*’ Variables with P-value $<0.05$ , Educ. level Education level, No formal educ. No formal education, Prim educ. Primary education, Sec. and above Secondary and above, Dist-(Km) Distance from the river in Km

### Performance of the Test Ov16 ELISA as a Reference for Ov16 RDT

The Ov16 ELISA test is expected to be to some degree more sensitive than skin snip microscopic detection in areas where onchocerciasis is hypoendemic or light infection situation, here of Ov16 ELISA test is used as reference for RDT and skin snip test. In this study, since skin snip microscopy is all negative the Ov16 ELISA and RDT are the compared tests. The test showed less sensitivity and higher specificity, indicated as 25.00% (9.77%-46.71%), 99.17% (97.02%-99.90%), 9.09% (5.91-13.22%), 75.00% (39.04%-93.36%), 92.97% (91.30%-94.34%) and sensitivity, specificity, disease prevalence, positive predictive value, and negative predictive value respectively.

## Discussion

Onchocerciasis remains a major public health NTD problem in developing countries especially, the Sub-Saharan African countries (21). Poor health facilities, health infrastructure, and health coverage are the major front-runner factors that made transmission of the disease is ongoing and protracted elimination. One major part of elimination program is mapping hypoendemic areas of the disease. The finding of this study showed that distance from the river, sex, village type, and age have significant association with positivity with Ov16 ELISA test. Communities residing nearer to the river showed seven times more likely positive than these reside far, males are nearly four times more positive than females for Ov16 ELISA test.

More than half 163 (61.7%) of the study participants were aware of the disease especially with the local name of “Foket” or “Yekolla ekek”; this might be due to the endemicity of the disease in the district and adjacent woreda’s. The worst number in this study was knowledge of the participants on etiology of the disease; they knew only 2.7%, which made this study not agreeable with other studies. Nearly half of this study knew that the mode of transmission of the disease is caused by blackfly biting, with at least one misconception like having contact with an infected person, house flies biting, mosquito biting, aerosol, and taking shower with dirty water. “Poor knowledge about the disease is a common factor in African rural inhabitant’s predisposing them to infection”. This current study is consistent, with studies done in North Ethiopia Quara focus and Southwestern Bebeka focus (22, 23).

In this study, about 9.1% of the study participants were positive for Ov16 ELISA test, 3% for Ov16 RDT test, and microscopic skin snip results were zero microfilaria for all participants of the study. This study is consistent with a study done in Northern and Central Togo after a decade of ivermectin mass drug administration, the sero-prevalence of the Ov16 ELISA test is 14.9% and the prevalence of the study done in Western Africa selected villages were <1% for Ov16 RDT test. Study done in Colombia Naiciona village for elimination verification showed the skin snip microscopic detection of microfilaria and Ov16 ELISA test for children ≤10 years old was zero; which is not in line with this study. The justification for this difference might be the elimination mapping in Naiciona focus was done after 22 rounds of ivermectin MDA conducted. But this study agrees with a study done in Equatorial Guinea Bioko Island focus with skin snip result of 0% and Ov16 sero-prevalence of 7.9% (20, 24-26). The value 9.1% Ov16 ELISA test is higher than other studies reported in Uganda Itwara focus which was 0% (27), Uganda Wadelai focus 0.03% (28), Sudan Abu Hamed focus 0% (29), Nigeria Nasarawa and Plateau focus 0.03% (30) and Ethiopia Metema Galabat focus 1.13% (31). The possible discrepancy might be explained, all those studies illustrated above were done after a year or a decades of ivermectin mass drug administration (MDA) in the focuses, yet the current study was done in the focus where MDA (ivermectin) was not administered.

The prevalence of morbidity indices for onchocerciasis of our finding was 6 (2.3%), 12 (4.5%) and 9 (3.4%) for palpable nodule, skin discoloration, and skin depigmentation respectively, which is not inconsistent with study done in Benishangul-Gumuz Asosa district with palpable nodule 30 (1.9%), skin discoloration 94 (6%) and skin depigmentation 38 (2.4%) and in SNNP Yeki district with the prevalence of palpable nodule 37 (2.6%), skin discoloration 33 (2.3%), 57 (4%) skin depigmentation (21).

In our finding, there is a significant association between sex and positivity for Ov16 ELISA test for onchocerciasis. Males were four times more likely positive for IG4 antibody recombinant to Ov16 antigen ELISA test than females. This study is in line with study done in Ethiopia Benchi Maji focus (21). This association could be due to males being more participants in outdoor/field activity especially in farming, fishing and irrigation.

This study illustrates that there is a significant association between participants age with positivity for IG4 antibody recombinant to Ov16 antigen ELISA test. The age group ranges from 21-41 17 (70.8%) years were nearly five times more likely positive than the other age category. This study agrees with a study done in Equatorial Guinea Bioko focus (20). The justification for the high rate among adults might be they are the productive age group participating in field activity; expected to be more exposed to black-fly biting. There was one (0.22%) positive from children less than 10 years old; meaning that IG4 antibody is detected in the child; which shows children were infected by the parasite, though the transmission the disease is ongoing in the community. Including school-age children under 10 in the study can maximize the positivity rate of the test, is a study unit which is not included under this study.

Samples collected from participants who reside in the first-line village were almost four times more likely positive for IgG4 antibody recombinant to Ov16 ELISA test than the random village residents. This association between the first-line and random village may be due to the first-line village residents more likely exposed to blackfly biting. Distance from the river is another variable that had an independent association with positivity with Ov16 ELISA test. Communities of the district living nearest to the fast-flowing river (<2Km) were seven times more likely 19 (79.1%) positive than those residing far (>5Km) from the river. Our study is consistent with a study done in Northern and Central Togo and Equatorial Guinea Bioko focus (20, 24). The association might be explained that communities who rely on their life on the river bases and fast-flowing rivers exposed to black-fly biting more frequently than who reside far.

The test performance showed poor sensitivity and a high specificity, 25.00% (CI 9.77%-46.71%) and 99.7% (CI 97.02%-99.90%) respectively. The study also displays the Positive Predictive Value 92.42% (CI 91.30%-94.34%) which is in line with research done in selected African and Latin American endemic zones (focus) (26). Having low sensitivity might be enlightened due to the small sample size of the study, so increasing the sample size may maximize the performance of the test.

## Strength of the Study

It includes the most important variables which have significant association with positivity for IgG4 antibody test against Ov16 recombinant antigens that can address and indicate the prevalence of onchocerciasis in the district; also the study included first-line villages that have high blackfly breeding, fast-flowing rivers with the communities living very nearest to the river; with high complaints of morbidity of the disease.

## Limitation of the Study

The first limitation of this study is serological studies didn’t differentiate current infections from the old one. Second, this study didn’t include school-age children which were better show the existence of active transmission in the community. Third, there may be a cross-reaction with a parasite Mansonella species which is an endemic in some Africa countries.

## Conclusion

In this study gender, village type, distance from the river and sex were the most significantly associated variables with positivity for Ov16 ELISA test and distance from the river was another variable significantly associated. In children under 10 years, one (0.4%) participant found positive for IgG4 antibody recombinant Ov16 antigen ELISA test, indicating an exposure of under 10 years children, so there might be ongoing transmission in the district. Communities that reside in the first-line village were more likely infected than communities residing in the random village. The serological test performance of this study was less sensitive and had profound specificity.

## Data Availability

All relevant data are within the manuscript

## Recommendation

Based on the findings of the study, the following recommendations are forwarded:

### To North Achefer health professionals

Giving health education to the residents of the district about onchocerciasis and its treatment, particular emphasis should be given for communities residing in the first-line village. Early diagnosis and management of morbidity indices in the community of the district should be given.

### To North Achefer district health bureau ad policymakers

Creating awareness, designing on the ways of controlling of the vector and increasing the accessibility of health care of the district is necessary.

### To future research

Researchers should increase the sample size to enhance the chance of finding positive for antibody or skin snip microfilaria and evaluating y molecular technique (i.e. PCR) and entomological techniques to maximize and ensure the exact status of the disease in the district.

## List of Abbreviations

APOC: African Programme for Onchocerciasis Control
CDTi: Community Directed Treatment with Ivermectin
DSB: Dried Blood Spot
ELISA: Enzyme Linked Immunosorbent Assay
EMO: Epidemiological Mapping of Onchorcerciasis
EOEEAC: Ethiopia Onchocerciasis Elimination Expert Advisory Committee
FMoH: Federal Ministry of Health
KAP: Knowledge, Attitude and Practice
MDA: Mass Drug Administration
NPV: Negative Predictive Value
NTD: Neglected Tropical Disease
OCP: Onchorcerciasis Control Program
PPV: Positive Predictive Value
RDT: Rapid Diagnostic Test
REMO: Rapid Epidemiological Mapping of Onchorcerciasis

## Ethical Consideration

Ethical clearance was obtained from the Research and Ethics Committee of the Department of Medical Microbiology, Immunology and Parasitology of AAU and the official letter was sent to Amhara Regional health office and to North Achefer district health bureau. After getting permission from the district to participate in the study, verbal and written consent obtained for the willingness of patients to participate. The participant’s identity was kept anonymously throughout the data collection and analysis period.

## Acknowledgments

Thanks to my advisors Dr. Tadesse Kebede and Mr. Sindew Mekasha for their unreserved support and guidance at each steps of my thesis. Next I would like to give my heartfelt thanks to AAU College of Health Science, Department of Medical Microbiology, Immunology and Parasitology, and Ethiopia Public Health Institute in general providing opportunity, their unfailing support and guidance in the study. I am also thankful to study participants, community leaders and health officials of North Achefer district.

## Author’s Information

Wuletaw Tadesse Mekonnin (BSc. Medical Laboratory Science) Assistance lecture at Salale University, MSc candidate by Medical Parasitology at Addis Ababa University; sponsored by Salale University, Fitche Ethiopia.

## Author’s Contribution

WT involved in conception and design of the study, supervised data collection, analysis and interpretation of data, and prepared the manuscript. TK and SM participated in the study design, reviewed the study instrument and data analysis, supervision, and involved in manuscript writing and editing.

## Availability of Data

In this manuscript all the data generated during the study included.

